# Rapidly emerging SARS-CoV-2 B.1.1.7 sub-lineage in the United States of America with spike protein D178H and membrane protein V70L mutations

**DOI:** 10.1101/2021.05.14.21257247

**Authors:** Lishuang Shen, Jennifer Dien Bard, Timothy J. Triche, Alexander R. Judkins, Jaclyn A. Biegel, Xiaowu Gai

**Author notes:** Correspondence to: Xiaowu Gai, PhD, Center for Personalized Medicine, Division of Genomic Medicine, Department of Pathology and Laboratory Medicine, 4650 Sunset Blvd., Mailstop #173, Los Angeles, CA 90027.

## Abstract

The SARS-CoV-2 B.1.1.7 lineage is highly infectious and as of April 2021 accounted for 92% of COVID-19 cases in Europe and 59% of COVID-19 cases in the U.S. It is defined by the N501Y mutation in the receptor binding domain (RBD) of the Spike (S) protein, and a few other mutations. These include two mutations in the N terminal domain (NTD) of the S protein, HV69-70del and Y144del (also known as Y145del due to the presence of tyrosine at both positions). We recently identified several emerging SARS-CoV-2 variants of concerns, characterized by Membrane (M) protein mutations, including I82T and V70L. We now identify a sub-lineage of B.1.1.7 that emerged through sequential acquisitions of M:V70L in November 2020 followed by a novel S:D178H mutation first observed in early February 2021. The percentage of B.1.1.7 isolates in the U.S. that belong to this sub-lineage increased from 0.15% in February 2021 to 1.8% in April 2021. To date this sub-lineage appears to be U.S.-specific with reported cases in 31 states, including Hawaii. As of April 2021 it constituted 36.8% of all B.1.1.7 isolates in Washington. Phylogenetic analysis and transmission inference with Nextstrain suggests this sub-lineage likely originated in either California or Washington. Structural analysis revealed that the S:D178H mutation is in the NTD of the S protein and close to two other signature mutations of B.1.1.7, HV69-70del and Y144del. It is surface exposed and may alter NTD tertiary configuration or accessibility, and thus has the potential to affect neutralization by NTD directed antibodies.

## Introduction

B.1.1.7 emerged in the UK and was the first major SARS-CoV-2 variant of concern (VOC) that is both more transmissible and apparently more virulent (1). It now accounts for 50-90% of the COVID-19 cases in U.S. and Europe. The Spike (S) protein N501Y mutation in the receptor-binding domain (RBD) confers higher binding affinity of the S protein for ACE2, while the other two deletions, HV69-70del and Y144del in the N-terminal domain (NTD) may also play a role in ACE2 receptor binding or neutralizing antibody escape (2). With millions of new B.1.1.7 cases in recent months, there is a very high probability of continuous acquisitions of new mutations, some of which may result in the emergence of new and even more infectious sub-lineages of B.1.1.7. While these new mutations may not be significantly deleterious by themselves, but when they appear in the context of other mutations within this VOC the result may be a more transmissible or pathogenic virus. This calls for rigorous genomic surveillance for newly acquired mutations in previously reported VOCs, including but not limited to B.1.1.7 and B.1.351.

Using the Children’s Hospital Los Angeles (CHLA) COVID-19 Analysis Research Database (CARD) (3), and viral sequences submitted to GISAID and NCBI GenBank, we have routinely performed genomic epidemiology and genomic surveillance studies of local, national and international databases (4–9). This allowed us to identify a new VOC (B.1.575) with a signature mutation I82T in the M gene (7). In the same study, we identified multiple other M mutations including V70L that are currently being encountered with significantly increased frequency. We have identified the M:V70L mutation in multiple SARS-CoV-2 lineages but primarily in the B.1.1.7 lineage. The B.1.1.7-M:V70L sub-lineage has been circulating at consistently moderate prevalence since November 2020. Through continuous genomic surveillance we identified the subsequent acquisition of yet another Spike mutation, D178H, within this lineage. This new B.1.1.7 sub-lineage, carrying both M:V70L and S:D178H mutations, appeared in February 2021 but by April 2021 quickly increased to account for 36.8% and 1.8% of all reported B.1.1.7 genomes in Washington and the U.S., respectively, This B.1.1.7-M:V70L-S:D178H sub-lineage is currently exclusive to the U.S. as of May 8, 2021, and had been detected in 31 states, with the majority of cases found in Washington, California and Ohio. Here, we report its detection, characterization, transmission, and evolution.

## Materials and methods

### Ethics approval

Study design conducted at Children’s Hospital Los Angeles was approved by the Institutional Review Board under IRB CHLA-16-00429.

### SARS-CoV-2 whole genome sequencing

Whole genome sequencing of the 2900 samples previously confirmed at Children’s Hospital Los Angeles to be positive for SARS-CoV-2 by reverse transcription-polymerase chain reaction (RT-PCR) was performed as previously described (5).

### SARS-CoV-2 sequence and variant analysis, and emerging variant monitoring

Full-length SARS-CoV-2 sequences had been periodically downloaded from GISAID (10, 11) and NCBI GenBank. They were combined with SARS-CoV-2 sequences from CHLA patients, annotated, and curated using a suite of bioinformatics tools, CHLA-CARD, as previously described (3). A custom Surging Mutation Monitor (SMM) standardized and integrated the viral genome and demographic data, in order to identify the trend of surging mutations and lineages across state and country levels. The current study was based on the 1.33 million global viral genomes that were available on May 1st, 2021.

### Phylogenetic analysis

Phylogenetic analysis was conducted using the NextStrain phylogenetic pipeline (version 3.0.1) (https://nextstrain.org/). Mafft (v7.4) was used in multiple sequence alignment (12), IQ-Tree (multicore version 2.1.1 COVID-edition) and TreeTime version 0.7.6 were used to infer and time-resolve evolutionary trees, and reconstruct ancestral sequences and mutations (13, 14). Phylogenetic analysis.

### Protein structure prediction

Structural predictions of mutant Spike proteins were carried out against the wild type protein PDB qhd43416 (https://zhanglab.ccmb.med.umich.edu/COVID-19/) using Missense3D service hosted online by the Imperial College London (http://www.sbg.bio.ic.ac.uk/~missense3d/) (15). CoV3D was used as the Spike Protein Mutation Viewer for the multiple mutations in B.1.1.7 (https://cov3d.ibbr.umd.edu/MutViewer/QTY83983).

## Results

### Identification of a rapidly emerging B.1.1.7 sub-lineage

We evaluated 1,333,679 SARS-CoV-2 viral genomes available on May 1^st^, 2021, including 2,900 from our own institution and the rest from GISAID and NCBI GenBank. We searched for SARS-CoV-2 mutations with significantly higher prevalence rate in both the U.S. and globally. Candidate mutations were further partitioned by pangolin lineage to identify emerging mutations in the context of a specific lineage, such as B.1.1.7 or B.1.351. We focused initially on the M mutations that we previously identified, including V70L, that were spiking near the end of 2020 (7). Overall the percentage of isolates that carried the M:V70L mutation had been relatively stable in the U.S. and globally with a gradual month to month increase (**Table 1**). In the vast majority of cases, the M:V70L mutation occurred on the B.1.1.7 lineage. While the percentage of B.1.1.7 isolates with the V70L mutation remained relatively stable across the world, the percentage fluctuated significantly in the U.S., attributable largely to the initial small number of B.1.1.7 cases in the U.S.

**Table 1.** **Number of isolates from the B.1.1.7 lineage and with the M:V70L mutation in the U.S. and globally**

| Month | Country | All isolates | B.1.1.7 isolates | All isolates w/ M:V70L | B.1.1.7 isolates w/ M:V70L | % all isolates w/ M:V70L | % B.1.1.7 isolates w/ M:V70L | % M:V70L in B.1.1.7 |
| --- | --- | --- | --- | --- | --- | --- | --- | --- |
| 2021-04 | U.S. | 44878 | 25837 | 720 | 716 | 1.60 | 2.77 | 99.44 |
| 2021-03 | U.S. | 91468 | 36034 | 965 | 953 | 1.06 | 2.64 | 98.76 |
| 2021-02 | U.S. | 56669 | 6859 | 309 | 287 | 0.55 | 4.18 | 92.88 |
| 2021-01 | U.S. | 52532 | 1365 | 193 | 150 | 0.37 | 10.99 | 77.72 |
| 2020-12 | U.S. | 23593 | 117 | 46 | 43 | 0.50 | 36.75 | 93.48 |
| 2020-11 | U.S. | 17878 | 2 | 4 | 1 | 0.02 | 50.00 | 25 |
| 2021-04 | World | 109886 | 82392 | 1257 | 1253 | 1.14 | 1.52 | 99.68 |
| 2021-03 | World | 291238 | 195853 | 2607 | 2592 | 0.90 | 1.32 | 99.42 |
| 2021-02 | World | 219853 | 107841 | 1845 | 1816 | 0.84 | 1.68 | 98.43 |
| 2021-01 | World | 195282 | 69514 | 1005 | 932 | 0.51 | 1.34 | 92.74 |
| 2020-12 | World | 107503 | 18356 | 257 | 236 | 0.24 | 1.29 | 91.83 |
| 2020-11 | World | 81587 | 2222 | 39 | 29 | 0.05 | 1.31 | 74.36 |

We identified the acquisition of another S mutation, D178H, in this B.1.1.7 sub-lineage (**Figure 1, Table 2**), which was estimated to have occurred on January 23, 2021. By April, the prevalence of SARS-CoV-2 isolates carrying the S:D178H mutation increased to 1.05% nationally and as high as 14.77% in Washington. When we examined the prevalence of S:D178H in the context of the B.1.1.7 lineage and the B.1.1.7-M:V70L sub-lineage, the numbers were even more striking (**Figure 2**). While the percentage of all B.1.1.7 isolates that carried the S:178H mutation increased from 0% in January 2021 to 1.8% in April 2021, there was a significant increase from 0% to 64.8% in B.1.1.7-M:V70L sub-lineage isolates that carried the S:D178H mutation were observed in the same time period. (**Figure 2A, Table 2**). The B.1.1.7-M:V70L-S:D178H sub-lineage was exclusive to the U.S. (**Table 2**), and findings when examined globally were not as striking (0% to 0.56% and 0% to 37%, respectively) (**Figure 2A, Table 2**).

**Table 2.** **Number of isolates form the B.1.1.7 lineage and with the S:D178H mutation in the U.S. and globally**

| Month | Country | All isolates | B.1.1.7 isolates | All isolates w/ S: D178H | B.1.1.7 isolates w/ S:D178H | % all isolates w/ S: D178H | %B.1.1.7 isolates w/ S:D178H | % S:D178H from B.1.1.7 isolates |
| --- | --- | --- | --- | --- | --- | --- | --- | --- |
| 2021-04 | U.S. | 44878 | 25837 | 469 | 464 | 1.05 | 1.80 | 98.93 |
| 2021-03 | U.S. | 91468 | 36034 | 402 | 399 | 0.44 | 1.11 | 99.25 |
| 2021-02 | U.S. | 56669 | 6859 | 18 | 10 | 0.03 | 0.15 | 55.56 |
| 2021-01 | U.S. | 52532 | 1365 | 5 | 0 | 0.01 | 0.00 | 0.00 |
| 2021-04 | World | 109886 | 82392 | 469 | 464 | 0.43 | 0.56 | 98.93 |
| 2021-03 | World | 291238 | 195853 | 403 | 400 | 0.14 | 0.20 | 99.26 |
| 2021-02 | World | 219853 | 107841 | 20 | 11 | 0.01 | 0.01 | 55.00 |
| 2021-01 | World | 195282 | 69514 | 7 | 0 | 7 | 0.00 | 0.00 |

**Figure 1.**
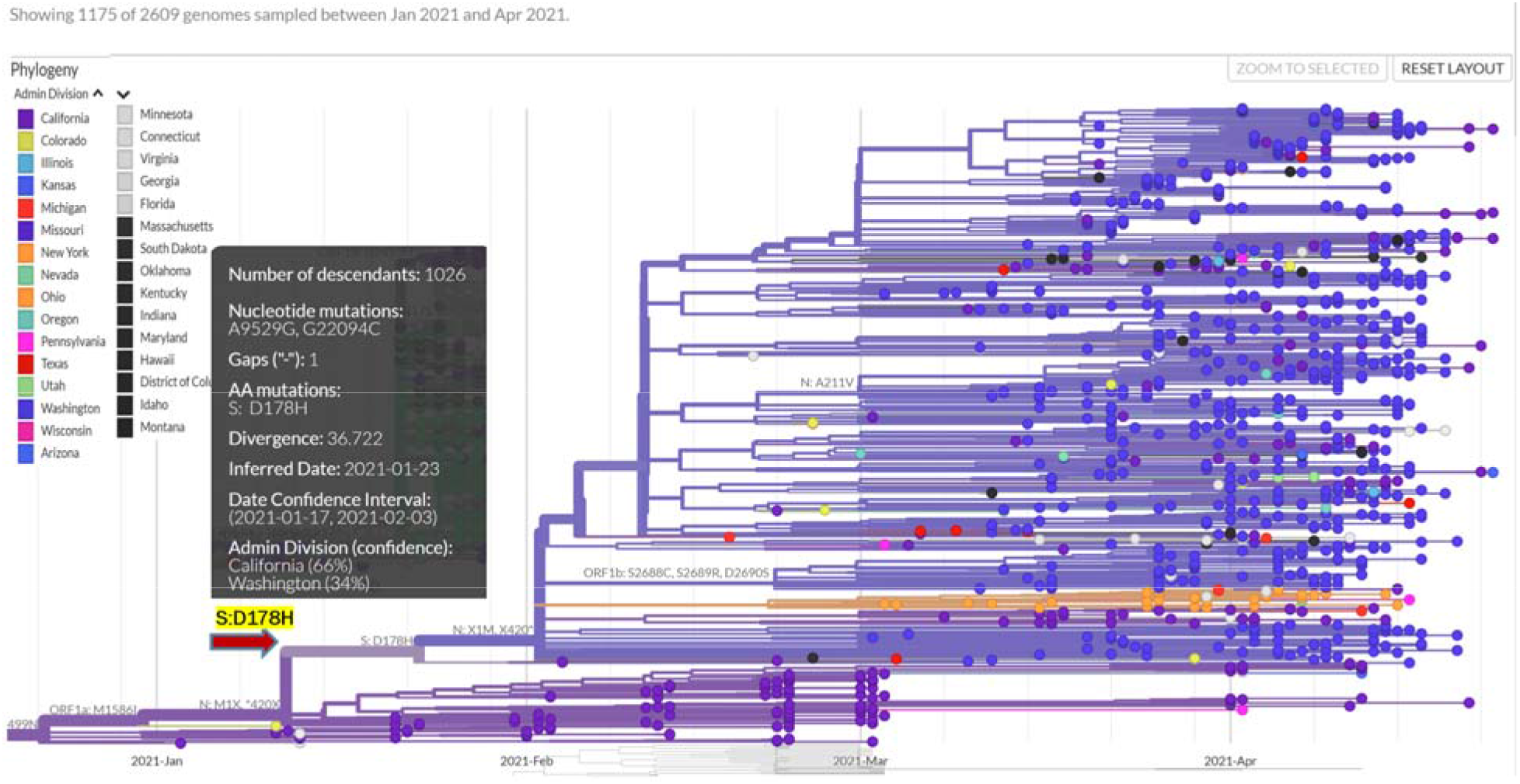
Branching off a sub-lineage of B.1.1.7 that is distinguished by the M:V40L mutation because of the emergence of the S:D178H mutation on January 23^rd^, 2021 as estimated. Each dot represents an isolate and is colored according to the state where it wa reported. The inset provides more information about the branching S:D178H mutation, namely the genomic position, the inferred date, the data confidence interval, and the likely states of origin.

**Figure 2.**
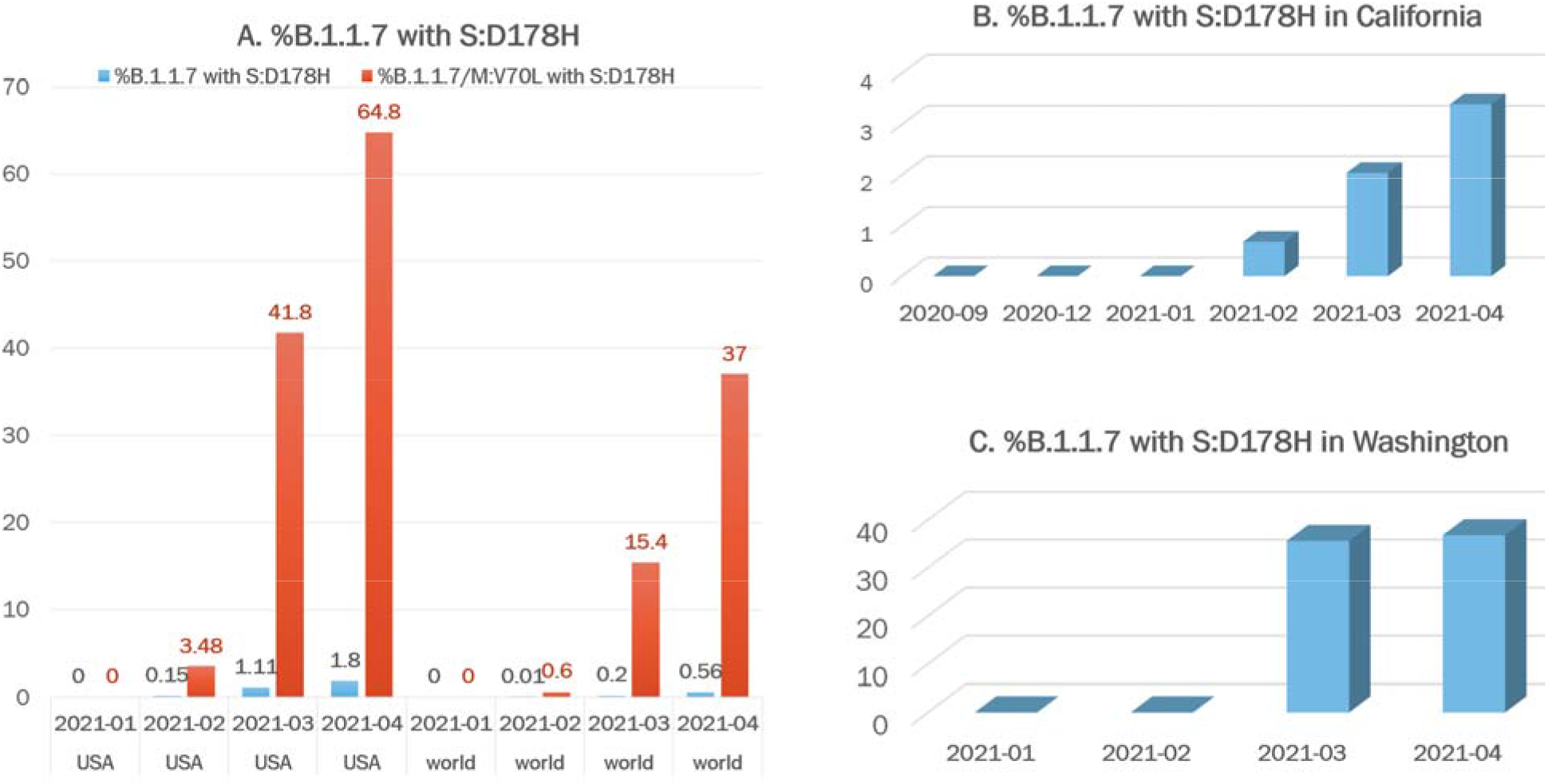
Prevalence of the B.1.1.7 sub-lineage carrying mutation S:D178H Globally, U.S., Washington, and California. A. Percentages of B.1.1.7 (blue) or B.1.1.7-M:V70L (orange) isolates that carried the S:D178H mutation from January to April 2021 in U.S. and globally, respectively. B. Percentages of B.1.1.7 isolates that carried the S:D178H mutation from September 2020 to April 2021 in California. C. Percentages of B.1.1.7 isolates that carried the S:D178H mutation from January 2021 to April 2021 in Washington.

The S:D178H mutation was also seen in other lineages including B.1.234 and B.1. In February, it was detected 10 times in 6 states and the first two were reported in California on February 4^th^, both within this B.1.1.7 sub-lineage. Overall, 98.93% of the S:D178H carrying viral isolates belonged to this new B.1.1.7 sub-lineage.

Within the U.S., most isolates of this lineage were from Washington, California, and Ohio (**Figure 3**). California and Washington showed the greatest increases in terms of the percentage of B.1.1.7 isolates carrying the S:D178H mutation. In Washington, the S:D178H mutation was absent until February 2021, where it quickly increased to account for 9.6% (296/3077) of all viral isolates in March and 14.8% (364/2464) in April (**Table 3**). It accounted for 36.8% (364/990) of the April B.1.1.7 isolates in Washington (**Figure 2C, Table 3**). In California, S:D178H was first seen in December 2020, but it was not seen within the B.1.1.7 lineage until February 4^th^, 2021 (**Table 3**). Its prevalence increased to 1.6% (45/2904) in April compared to all isolates studied in California, and 3.3% (45/1353) of the B.1.1.7 isolates (**Figure 2C, Table 3**).

**Table 3.** **Number of isolates from the B.1.1.7 lineage and with the S:D178H mutation in the states of California and Washington**

| State | Month | All isolates | B.1.1.7 isolates | Isolates w/ S:D178H | % of all isolates w/ S:D178H | % B.1.1.7 isolates w/ S:D178H |
| --- | --- | --- | --- | --- | --- | --- |
| California | 2021-04 | 2904 | 1353 | 45 | 1.55 | 3.33% |
| California | 2021-03 | 9951 | 2254 | 46 | 0.46 | 2.04% |
| California | 2021-02 | 8889 | 589 | 10 | 0.11 | 0.68% |
| California | 2021-01 | 10441 | 214 | 4 | 0.04 | 0% |
| Washington | 2021-04 | 2464 | 990 | 364 | 14.77 | 36.8% |
| Washington | 2021-03 | 3077 | 832 | 296 | 9.62 | 35.6% |
| Washington | 2021-02 | 2152 | 115 | 0 | 0.0 | 0% |
| Washington | 2021-01 | 1510 | 33 | 0 | 0.0 | 0% |

**Figure 3.**
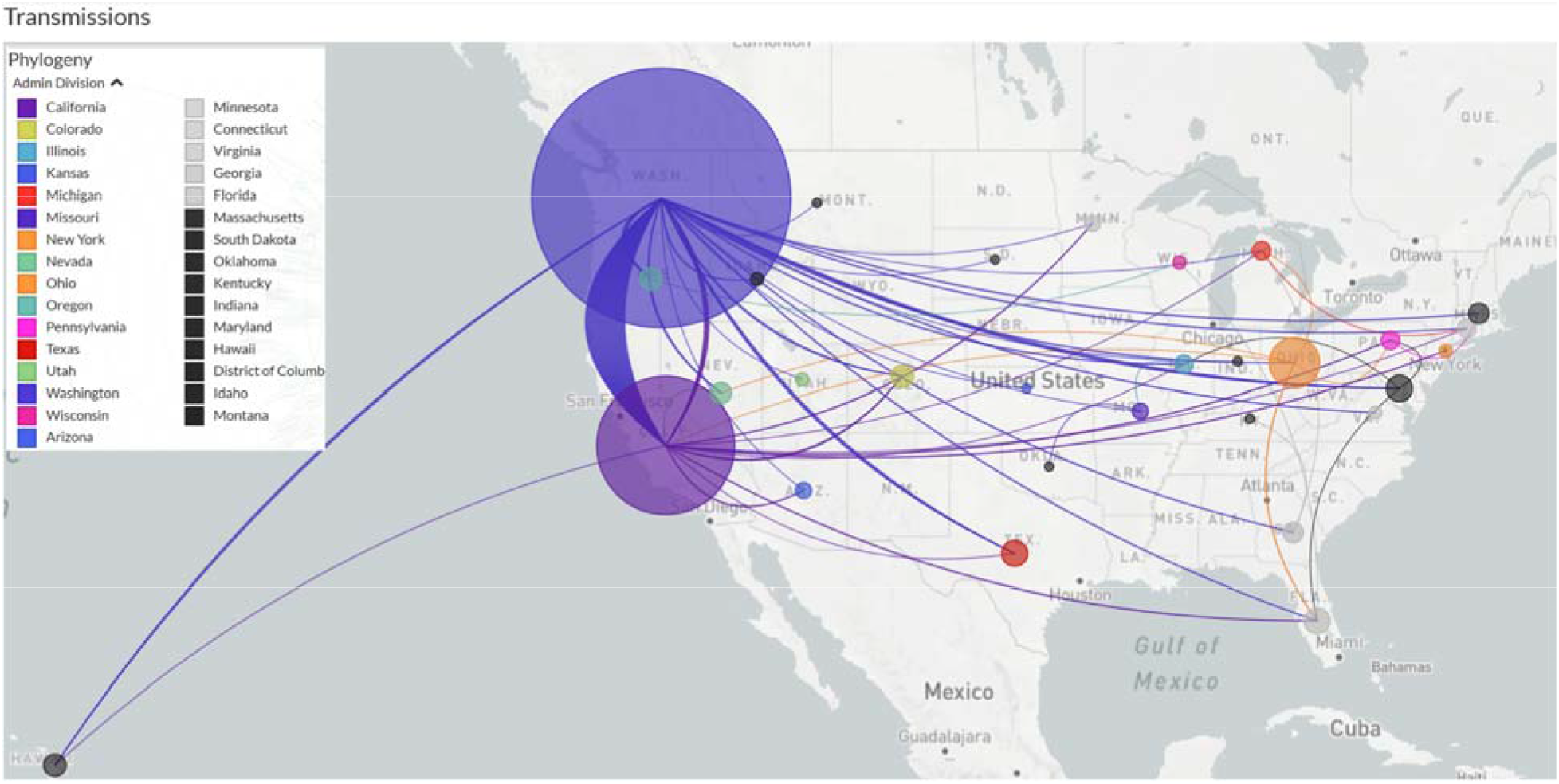
Transmission of the 1026 B.1.1.7-MV70L-S:D178H isolates across the U.S.. The size of the circles are drawn proportional to the number of cases in the specific state. Lines are colored the same as the exporting states.

### Origin and transmission of the B.1.1.7-M:V70L-S:D178H sub-lineage

To understand the origin and evolution of this B.1.1.7-M:V70L-S:D178H sub-lineage, we used a) all 1,125 S:D178H carrying viral sequences reported across the globe, b) 1,600 subsampled M:V70L carrying viral sequences across the world, and c) the NC_045512 reference genome. These sequences were further filtered with requirements of being at least 27,000 bp and having complete date information. 2,609 genomes were kept for final phylogenetic and transmission analysis using Nextstrain, rooted by NC_045512 (**Figure 1**). The D178H branch divergence date was estimated to be January 23, 2021, with a date confidence interval of January 17, 2021 to February 3, 2021. It likely originated in California (66% probability) or Washington (34% probability).

Estimated transmission routes are demonstrated in **Figure 3**, where the size of the circle represents the number of cases from this sub-lineage in each state, and the line colors correspond to the exporting locations, California, Washington, and Ohio contributed the majority of transmissions compared to other states. This sub-lineage was clearly exclusive to the U.S., and not reported in any other countries at the time of the study.

### Signature mutations of the B.1.1.7-M:V70L-S:D178H sub-lineage

Within this sub-lineage, the most common signature mutations were the same as B.1.1.7 signature mutations, with the additional M:V70L and S:D178H mutations. There were no other novel common spike mutations within this sub-lineage.

### Protein structure and mutation effect prediction

The 3D structure of the Spike protein, as visualized using the CoV3D mutation viewer. Using these results we were able to show that the S:D178H mutation is close structurally to two signature deletions of B.1.1.7, HV69_70del and Y144del (**Figure 4**). They are all surface exposed and likely alter N terminal domain (NTD) tertiary configuration.

**Figure 4.**
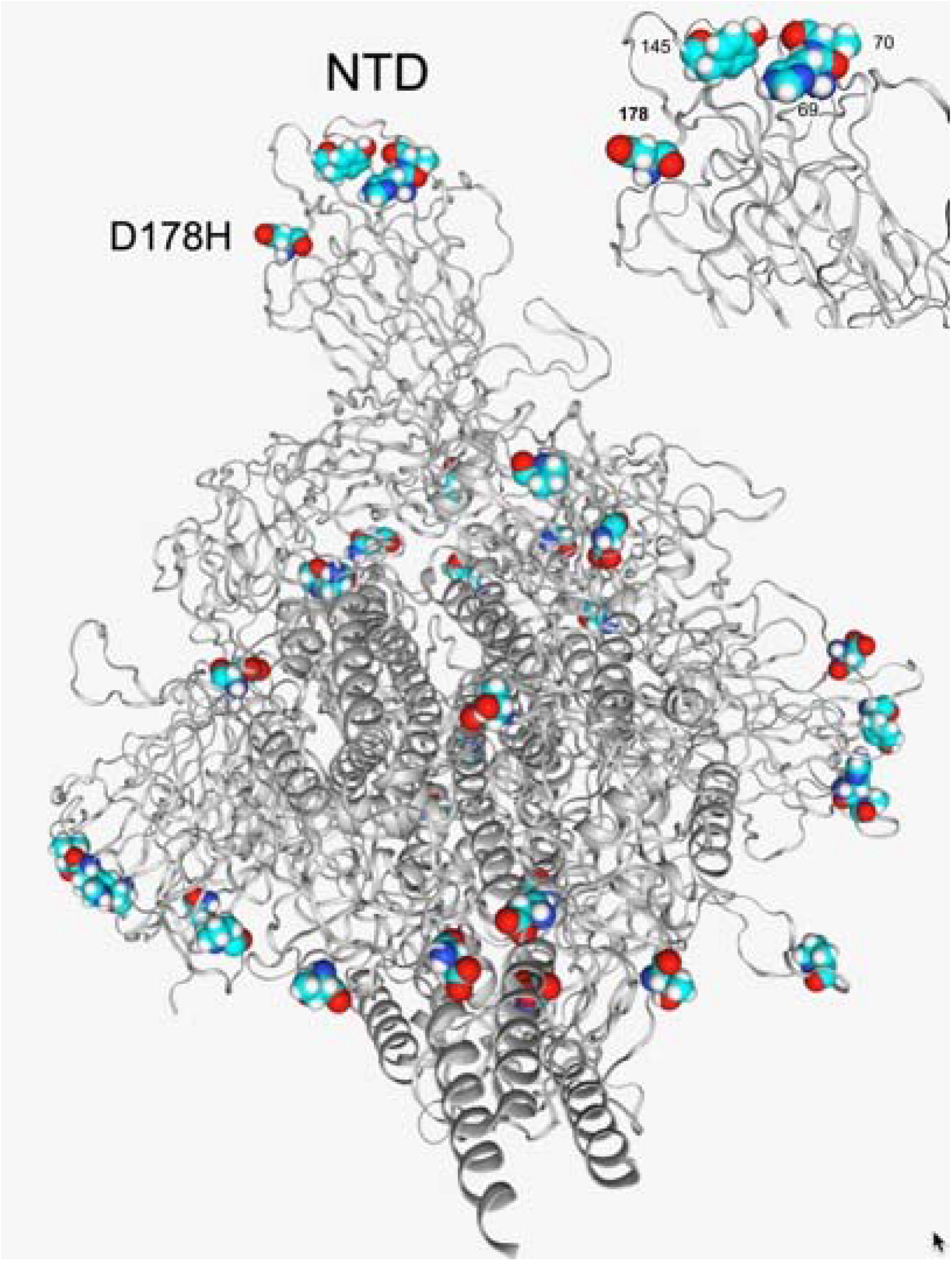
Structural location of the D178H and other spike protein mutations in the B.1.1.7 lineage. D178H is in close proximity to other amino acids in the NTD that are affected in the B.1.1.7 lineage, HV69-70del and Y144/Y145del (top right).

## Discussion

In March 2021, the B.1.1.7-M:V70L-S:D178H lineage appeared abruptly at high prevalence (35.6%) in all reported B.1.1.7 isolates in Washington. This was a remarkable finding as it was absenting in 115 Washington B.1.1.7 isolates reported in February. This abrupt change could reflect undersampling or potentially reflect superspreader events. Given the spike in the number of B.1.1.7 cases, this sub-lineage clearly appears to be more transmissible than even the original B.1.1.7 lineage given the spike in the number of B.1.1.7 cases. This finding warrants prompt and further attention by public health authorities, as this mutation profile is closely linked to the resurgence of cases in Washington in particular. It also strongly supports the now widely recognized need for more extensive SARS-CoV-2 viral sequencing of PCR positive COVID-19 cases for detection of new mutations of concern as part of widespread genomic surveillance (16, 17).

The S:D178H mutation, while demonstrably associated here with the more pathogenic B.1.1.7 lineage, is not necessarily by itself more pathogenic. Dozens of SARS-CoV-2 genomes that carry the S:D178H mutation were reported before February, but none of these demonstrated the increased frequency seen when the mutation occurs in the context of the B.1.1.7 lineage. Phylogenetic analysis revealed a distinct and long branch leading to the new S:D178 branch after M:V70L. Together, these observations suggest that the S:D178H mutation is recurrent, but only increased exponentially in the context of the more pathogenic B.1.1.7 lineage, which serves as an argument for its fitness. This is the same observed with other deleterious mutations like the N501Y and E484K mutations, both of which are now superimposed on distinct and separate more pathogenic lineages. The S:D178H mutation arose independently again in the U.S. on the B.1.1.7-M:V70L background. The rapid increase in its prevalence, only after its acquisition by the B.1.1.7-M:V70L sub-lineage suggests this combination of mutations is associated with increased transmissibility. It is also of interest that this mutation occurs in the NTD, unlike most of the mutations associated with current VOC that are centered on the spike protein RBD, implying that NTD mutations beyond the original 69-70del and the 144del are of concern. And finally, it should be noted that this NTD mutation co-exists with the previously reported M protein mutation M:V70L, suggesting that M protein mutations also contribute to enhanced biologic ‘fitness’ or pathogenicity of this sub-lineage.

The appearance of the S:D178H mutation in the context of the B.1.1.7 lineage is temporally associated with the increased incidence of COVID-19 in Washington. New cases in Washington were higher than the national level at the time of the study. According to New York Time COVID-19 dashboard, the 7-day average of new cases on May 2 was 1,379 in Washington, which was only a 50% reduction compared to 2,757 cases on December 15. In comparison, the numbers on May 2 was 49,270 in U.S., a 77.3% reduction from the December 15 number of 21,7325. The appearance of this new B.1.1.7 sub-lineage temporally linked to increased cases in Washington warrants further investigation.

The potential effect of the S:D178H mutation on immunity and vaccine ‘escape’ also warrant further analysis. Mutations in the Spike N-terminal domain have been associated with lack of neutralization by NTD directed antibodies, especially when the N5 loop is affected (18, 19). The NTD initiates viral binding to the ACE2 receptor expressing host cell. Since the D178H falls in the NTD domain close to the N5 loop, it may alter NTD structure and antibody recognition. It may thus have a similar immune evasion effect as the HV69-70del and Y144del mutations, or it may further enhance that of the two other mutations, based on the 3D model. These findings highlight the continued importance of active genomic surveillance to monitor the spread of this B.1.1.7-M:V70L-S:178H lineage.

## Data Availability

The SARS-CoV-2 sequences used in the study were obtained from GISAID and NCBI GenBank. Additionally, we included in our study SARS-CoV-2 sequences that we obtained from our own institution's patients, but that were also submitted to GISAID.

## Acknowledgments

We would like to acknowledge all members of the Department of Pathology and Laboratory Medicine (PLM) at Children’s Hospital Los Angeles for dedication towards providing excellent patient care throughout the pandemic, including especially the rapid launch of both the COVID-19 diagnostic test in the Clinical Microbiology and Virology Laboratory and the SARS-CoV-2 whole genome sequencing assay at the Center for Personalized Medicine in March 2020, both with the support of Thermo Fisher and Paragon Genomics, with whom these assays were developed. We would like to acknowledge the frontline healthcare workers who remain devoted in the fight against COVID-19. We would also like to acknowledge all institutions that have contributed the SARS-CoV-2 sequences timely, as well as NCBI, GISAID, and Nextstrain for providing valuable resources for SARS-CoV-2 genomics.

